# Subcutaneous injections of penicillin (SCIP): Convenient and effective treatment for Māori, Pacific Peoples and their whānau in preventing rheumatic heart disease

**DOI:** 10.1101/2025.03.13.25323800

**Authors:** Julie Cooper, Monleigh Muliaumasealii, Dhevindri Moodley, Jacqui Ulugia, Anneka Anderson, Julie Bennett

**Affiliations:** Department of Public Health, University of Otago, Wellington, New Zealand; National Hauora Coalition, Auckland, New Zealand; Te Whatu Ora Capital, Coast and Hutt Valley, Wellington, New Zealand; Faculty of Medical and Health Sciences, Te Kupenga Hauora Māori, University of Auckland, Auckland, New Zealand

## Abstract

**Background:** Acute rheumatic fever (ARF) and rheumatic heart disease (RHD) remain significant health issues for Māori and Pacific communities in Aotearoa New Zealand (NZ). Subcutaneous injection of penicillin (SCIP) enables injections to be given 10-weekly as an alternative to the standard secondary prophylaxis of four-weekly intramuscular (IM) injections. As part of a clinical trial involving participants who have been on SCIP for at least one year, we aim to explore treatment adherence, pain management, and quality of life for Māori and Pacific participants and their whānau.

**Methodology:** A community centred approach aligned with Kaupapa Māori, and Pacific-centred research values was used. Semi-structured interviews were conducted with 10 whānau (family), including nine participants on SCIP. Data collection occurred between March and September 2024. Thematic analysis was used to identify key themes from participants’ experiences.

**Principal findings:** Six themes emerged: Reduced burden of treatment; emotional impact from reduced injection frequency; whānau-centered care by healthcare providers, relationship building (whakawhanaungatanga), health literacy, and pain management. Participants reported significant improvements in quality of life and valued SCIP’s flexibility, allowing injections at home, work, or school, with the added convenience of visits every 10-weeks. Strong relationships with healthcare providers, especially research nurses, were essential for adherence. Emotional barriers, such as stress associated with frequent injections, were reduced with SCIP.

**Conclusions:** This study highlights the benefits of SCIP in reducing physical and emotional barriers, which in turn enhances the quality of life for Māori and Pacific participants with ARF. The findings emphasise the importance of culturally responsive, whānau-centered models of care. SCIP has the potential to improve adherence to secondary prophylaxis, with the potential to be offered as a standard treatment, improving health outcomes across NZ and internationally.

## Introduction

Acute rheumatic fever (ARF) is a preventable inflammatory disease that occurs as a delayed sequelae to group A streptococcus (GAS) infection. ARF and its complication rheumatic heart disease (RHD) have significant negative effects on health, often resulting in chronic illness and premature death. It is estimated that 40.5 million people worldwide live with RHD, resulting in approximately 400,000 deaths each year [1, 2]. While ARF and RHD have nearly been eradicated in high-income nations, these conditions continue to cause significant, preventable harm and mortality in Aotearoa New Zealand (NZ), disproportionately affecting Māori and Pacific Peoples. [3].

For 70 years, the only proven way to prevent ARF progression has been benzathine penicillin G (BPG), which in NZ is given as a monthly intramuscular (IM) injection for a minimum of 10 years [4]. The effectiveness of this approach is limited by complex issues including the age of young patients and health system failures. Furthermore, pain, fear, and the frequency of injection are recognised as barriers which can lead to suboptimal adherence [5].

Recent work investigating the potential benefits of subcutaneous (SC) delivery of BPG has shown that the pharmacokinetic profile and tolerability of BPG delivered via SC delivery show a prolonged duration of effect, comparable pain scores, and equivalent safety [6]. Furthermore, the safety, tolerability, and pharmacokinetics of subcutaneous injection of penicillin (SCIP) was assessed in 24 healthy adult volunteers assigned to receive infusions of 3.6, 7.2, or 10.8 MIU (three, six, and nine times the standard dose, respectively), found that higher doses were safe, well-tolerated, and potentially suitable for up to three monthly dosing intervals for secondary prophylaxis of RHD [7]. A follow-on Phase II clinical trial of young people living with ARF who were given high-dose SCIP reported that participants preferred SCIP over their usual monthly IM penicillin regimen, citing less pain and a preference for the longer time gap, 70 versus 28 days, between treatments [8]. Reducing injection frequency from 13 to four-or-five per year, may improve adherence and reduce disease progression. This study builds on that work and aims to understand the lived experiences of participants who have been on SCIP for more than 12 months or received at least four SCIP injections, and the impact this has on them and their whānau.

## Methods

We used qualitative methodologies aligned to Kaupapa Māori and Pacific-centred research values [9]. This culturally aligned approach was critical, involving a senior researcher who identified as Māori, another researcher who identified as Pacific, and the field researcher conducting the interviews identified as Pākehā (non-Māori NZer). The Kaupapa Māori approach ensured the research respected Te Ao Māori (the Māori worldview) and mātauranga Māori (Māori knowledge), to ensure findings were meaningful and culturally relevant [10]. Similarly, the research respected Pacific cultural values, practices, and knowledge systems. The study was designed to benefit Pacific and Māori communities, with Pacific and Māori researchers reviewing and validating the approach to ensure accuracy and alignment with cultural values. All participants (or their parent / legal guardian) provided written, informed consent, witnessed by a study nurse/researcher, prior to participating. Consent was obtained for children under the age of 16 years. Information and consent forms were available in English, Te Reo Māori and Samoan.

### Participant recruitment

Tamariki (children) and rangatahi (youth) who had been on SCIP in the Wellington region for at least 12 months or received four or more SCIP injections were invited by a research nurse to participate during their regular SCIP appointment. The participants whānau (family) were also invited to share their experiences, recognising the central role of whānau in health, with wellbeing viewed as a collective responsibility. If SCIP participants or their whānau agreed to take part, the field researcher made contact to arrange an interview. Interviews were arranged at participants convenience with options of phone, Zoom, or in-person. In-person locations were chosen by participants to ensure participants felt culturally safe and included their homes, community spaces and cafes.

### Data collection

Semi-structured interviews were held in-person. Ethics covered the wider study which commenced on 21^st^ November 2022 with interviews with participants and their whānau, between 19^th^ March and 10^th^ September 2024. The key areas of focus were participants’ likes and dislikes of SCIP, its impact on their ability to engage in activities such as school, work, sports, and holidays, and comparing adherence to SCIP with that of IM BPG. Additionally, the study explored the impact of SCIP on whānau and their overall experience with the treatment. After ten interviews data saturation was reached with no new themes emerging. Interviews were auto recorded and lasted between 20-30 minutes. All interviews were transcribed verbatim by the field researcher.

### Data analysis

A general inductive thematic analysis was used to identify and categorise themes relevant to the research objectives [11]. The thematic analysis followed a process of data immersion, coding, categorising and generation of themes aligning with the theoretical approach described by Green and colleagues [12]. The field researcher (JC) engaged in deep immersion with the data to ensure a thorough understanding of participants’ experiences and perspectives. This involved repeatedly reviewing the transcripts and ensuring that the analysis remained grounded in whānau experiences. Through collaborative discussions, a coding framework was developed based on recurring keywords and phrases within the transcripts. Manual coding was conducted independently by the field researcher, the senior Māori researcher (AA), and the Pacific researcher (MM). The researchers then collaboratively grouped related codes into broader categories. These categories were developed through discussions that focused on capturing the essence of participants’ experiences, aligning with the research objectives and the respective cultural frameworks. The final step of the analysis was the identification of themes from the categories, where the three researchers analysed the categories and associated narrative data to identify patterns. The initial themes were reviewed by the broader research team and a Pacific participant with lived SCIP experience. Their input helped refine the themes to reflect Māori and Pacific cultural values and the lived experiences of participants more accurately, enhancing the study’s credibility and cultural relevance.

### Ethics approval and consent

Ethical approval was obtained from the NZ Health and Disability Ethics Committee (approval number 11094) and was endorsed by Te Whatu Ora, Capital Coast, and Hutt Valley Health, including a review by their Māori Research Board (RAG-M #916). Research consultation was undertaken with Ngāi Tahu Research Consultation Committee. Participant information sheets explained the study’s purpose and the types of questions that would be asked. Written informed consent/assent was obtained from all participants/guardians. Findings were shared with whānau and communities involved in the study.

## Results

### Characteristics of participants

Ten interviews were conducted. This included interviews with nine participants who had lived experience of SCIP, and six interviews with parents of children who were on SCIP. The mean age of SCIP participants was 17 years (range 13-23 years). Half of SCIP participants were male, six identified as Māori and four as Pacific (Samoan, Fijian, Cook Island Māori) See Table 1.

**Table 1:** Characteristics of participants.

| Whānau | Name<br>(Pseudonyms) | Participant | Ethnicity | Age Band<br>of SCIP<br>participant<br>(years) | Gender<br>of SCIP<br>participant |
| --- | --- | --- | --- | --- | --- |
| 1 | Wiremu | Patient | Māori | 15-19 | Male |
| 2 | Hine | Patient | Māori | 20-24 | Female |
| 3 | Maia<br>Tui | Patient<br>Mother | Māori<br>Māori | 15-19 | Female |
| 4 | Teuila | Patient | Cook<br>Islands | 15-19 | Female |
| 5 | Leilani<br>Mele | Patient<br>Mother | Samoan<br>Samoan | 15-19 | Female |
| 6 | Aroha<br>Marama | Patient<br>Mother | Māori<br>Māori | 10-14 | Female |
| 7 | Sione | Patient | Samoan | 15-19 | Male |
| 8 | Kiri | Mother | Māori | 10-15 | Male |
| 9 | Sevu<br>Mere | Patient<br>Mother | Fijian<br>Fijian | 10-15 | Male |
| 10 | Ari<br>Moana | Patient<br>Mother | Māori<br>Māori | 15-19 | Male |

### Thematic analysis

The thematic analysis identified six themes: *Reduced burden of treatment; emotional impact from reduced injection frequency; wh*ā*nau-centered care by healthcare providers, Whakawhanaungatanga (relationship building), health literacy, and pain management.* Themes and subthemes are summarised in Table 2.

**Table 2.** Summary of themes and subthemes for participants on subcutaneous injections of penicillin and their whānau.

| Theme | Subthemes |
| --- | --- |
| ***Reduced burden of treatment***<br>Reducing the frequency of injections offered greater convenience, enabling SCIP participants and their whānau to engage more easily in school, work, sports and holidays. | Supporting active participation in life activities<br>Reduced disruptions to daily living |
| ***Emotional impact from reduced injection frequency***<br>The emotional effects of treatment on both participants and whānau, focusing on reducing stress and increasing confidence in the care process. | Participant's impact<br>Whānau impact |
| ***Whānau-centered care by healthcare providers***<br>Placing the well-being of the entire whānau at the forefront, ensuring that healthcare is flexible, convenient, and minimally disruptive to daily life. | Adapting care to whānau contexts |
| ***Whakawhanaungatanga (relationship building)***<br>The importance of building strong, trusting relationships between participants, whānau and health care professionals developing a sense of belonging and mutual respect. | Making and maintaining relationships<br>Empowering through communication |
| ***Health literacy***<br>Empowering participants through continuous education and support, helping them better understand their health and navigate healthcare systems. | Informed self-care<br>Ongoing support and communication |
| ***Pain management***<br>The varying levels of pain participants experience with different treatment methods, highlights the trade-off between experiencing occasional discomfort with SCIP versus the more frequent pain associated with IM injections. | Differing levels of pain for participants<br>Pain trade-off with fewer injections |

### Reduced burden of treatment

Reducing the frequency of injections with SCIP offered greater convenience, allowing participants and their whānau to more easily engage in school, work, sports, and holidays. This reduced injection schedule minimised disruptions to daily life, empowering whānau to manage care in a way that supported their collective well-being. Participants reported fewer interruptions to their routines, leading to a more balanced and less stressful lifestyle. This is reflected below by mothers Mele, Marama, and Kiri.

> *“Every time she has her injection; it’s a big thing for me. I have to remember to make sure that she is not doing anything heavy, make sure she has to go have a rest. Since she’s on the three-monthly, it’s less stress for me as well.” Mele (mother)*
>
> *“I don’t see her go through pain every single month I can just see her go through pain once and it lasts a long time.” Marama (mother)*
>
> *“The convenience, and it’s only what four times a year rather than 12.” Kiri (mother)*

The most common comment from SCIP participants was the convenience of having fewer injections as described by Moana and her son Wiremu:

> *“You break it down four a year rather than 12.” Moana*
>
> *“Would you rather be chased up every month or be seen four times a year? I choose four times per year.” Wiremu*

### Fewer injections supported greater participation in day-to-day activities

A subtheme of reduced burden of treatment was the ability for greater participation in day-to-day activities. Participants were able to engage more in activities such as school, sports, and social events, thanks to the fewer and less disruptive injections. This flexibility was particularly important for children and young people who were balancing treatment with active lifestyles.

> *“We could have a long holiday away from here, six weeks in Samoa. This three-monthly injection was very helpful.” Mele (mother)*
>
> *He can just do more things, play rugby, helping around the home. Mere (mother)*

For participants on SCIP, it significantly eased the management of their own schedules. Switching from IM to SCIP has resulted in notable improvements in Teulia’s quality of life, as she has been on SCIP for two years. For example, she completed her first year of full-time nursing studies, took on two part-time jobs, and travelled abroad to the Cook Islands. Reflecting on the potential return to monthly injections, she expressed:

> *“It would be scary going back to it. I remember how bad it used to be… it feels like too much after getting used to SCIP.” Teulia*

### Reduced disruptions to daily living

Fewer injections with SCIP allowed participants to avoid the frequent interruptions caused by monthly injections. This reduction in treatment-related disruptions provided participants and their families more freedom in planning daily activities, resulting in a smoother and less stressful routine, as described by mother Tui.

> *“She does not take any time away from school, can go back to playing basketball and waka ama traditional Canoe paddling the next day. Is going to Samoa on a school trip, she is able to do this as her health is now much better and she is in a routine with the three-monthly injections.” Tui (mother)*

### Emotional impact from reduced injection frequency

The second theme that emerged was the emotional well-being of participants and their whānau, which improved significantly with SCIP, due to the reduced frequency and flexibility of injections. Both participants and caregivers experienced less anxiety and stress associated with treatment.

### SCIP participants

Participants reported a reduced emotional toll, the reduced frequency and flexibility of SCIP treatment resulted in less stress, anxiety, and mental strain compared to the monthly IM injections. Hine, Teuila and Wiremu’s comments were a very common theme from all of the participants.

> *“It’s literally not on my mind until I get the message. I have three months where I focus on other aspects of my life, work, home.” Hine*
>
> *“It’s a lot less to worry about on top of everything else.” Teuila*
>
> *“It does impact your mental health; you are not focused on that needle every month. You have more time to think to yourself.” Wiremu*

### SCIP participants wh***ā***nau

Caregivers also felt less stressed not having to worry about their child experiencing pain or remembering the monthly injection schedule. This flexibility allowed whānau to plan holidays and enjoy time together without the constant concern of treatment, leading to less emotional burden and fewer disruptions to their lives.

> *Everything is going smoothly moving from the monthly to the three monthly…it’s way less stress for us.” Mele (mother)*

SCIP participants Teuila and Hine now manage their own health care plans, with SCIP their whānau worry a lot less as the schedule is easier to manage.

> *“My mum’s happier because I missed a lot of my monthly injections once I swapped over into the adult sector… She does not have to worry about it.” Teuila*
>
> *“I couldn’t really do much overnight extracurriculars. If I didn’t get my shots because my mum would be stressed if I missed it by one day.” Hine*

### Wh**ā**nau-centered care by healthcare providers

The third theme that was identified was the well-being of the entire whānau was at the forefront, ensuring that appointments were flexible, convenient, and minimally disruptive to daily life.

### Adapting care to wh***ā***nau contexts

The nurses delivering SCIP allowed for flexible scheduling and location, adapting to whānau contexts and routines. This allowed participants to receive treatment in ways that best suited their routines. Participants appreciated the ability to choose injection times and locations that were most convenient for them and their families, as described below:

> *“They [research nurse] always text me in advance. They go to school during school time, and they come [to my] home during the holidays.” Mele (mother)*
>
> *“With the monthly injection it was either this day or this day and they [nurse] didn’t give me an option on the time so even if it was in the middle of my course or school, I just had to do it anyway.” Teuila*
>
> *“I have it [SCIP] at home as I don’t like hospitals, I spent too much time there (when diagnosed) and I don’t want to go back.” Wiremu*

### Whakawhanaungatanga (relationship building)

The fourth theme was Whakawhanaungatanga, which refers to connecting and getting to know people in their health care. Whakawhanaungatanga was integral in building trust and developing strong connections through open communication between healthcare providers, particularly between the Research Nurses and whānau. Whakawhanaungatanga created a supportive and collaborative environment that encouraged participants to transition from monthly IM to SCIP and improve adherence.

### Making and retaining relationships

Strong relationships, especially with the research nurses, enabled trust to be built and gave participants confidence in the SCIP procedure. This trust facilitated participants’ willingness to switch from IM to SCIP and improved adherence to the SCIP treatment. Having a constant point of contact, helped build trust and made SCIP participants feel more comfortable reaching out, which in turn enhanced their adherence.

> *“If it had not been for [research nurse], I would still be doing the monthly. We would talk at length about the pros and cons. She helped me understand more about it” Hine*

Teuila developed a strong relationship with the research nurse after some challenging experiences with her previous healthcare providers, when she had a difficult time transitioning to adult care. Her previous inconsistent and inflexible care lead to her missing her monthly injections. However, her experience with SCIP has been markedly different.

> *“Now I have a good relationship with the nurse, so I do not feel like I am getting the procedure. We just talk the whole time it’s getting done.” Teuila*

For some whānau it took several conversations with health care providers to gain reassurance that SCIP was the right option for their child. Like Kiri and Mele, many participants and whānau expressed anxiety and scepticism about transitioning to SCIP, particularly regarding the ‘trial’. It took time and repeated conversations with healthcare providers to alleviate these concerns.

> *“I was a bit sceptical of any medicines after Covid and about being part of a trial. It took a few times to agree, I think it was about a year before we said that would be cool.” Kiri*
>
> *“I declined the first letter because I was not sure, it wasn’t rolled out to the public. They were only selecting a few people to for trial.” Mele (mother)*

### Empowering through communication

Clear and open communication from research nurses helped participants understand the benefits of SCIP, alleviating their anxieties and empowering them to engage more actively in their care. It was particularly important for parents like Moana to feel comfortable with a new type of treatment.

> *“For people like me who are a bit weary when it comes to their children, about the introduction of new medicine, it seemed relatively safe, and we got enough information that I was comfortable.” Moana (mother)*

### Health literacy

The fifth theme that emerged was health literacy. Participants reported that by being part of the SCIP trial they gained a better understanding of what ARF was and how the treatment could prevent GAS infections, which could lead to disease progression. This knowledge empowered participants and their whānau to make informed decisions regarding SCIP and enabled them to better manage their health care.

### Informed self-care

A subtheme of health literacy was informed self-care. Having a greater understanding of what ARF was and the importance of self-care enhanced the participants’ ability to manage their treatment. Educating participants and their whānau increased their knowledge about their condition, treatment options and management. Like Hine, many of the participants and whānau developed a better understanding of what self-care they needed in the days following their injection.

> *“I didn’t take the advice seriously after the first injection. I went straight back to work. Like fully back into work. I wrecked myself, and the bruising went right across my stomach. It is way better now I know what to expect. The main pain is the first day. The bruising isn’t as bad now that I know not to constantly knock into it….I am able to do light duties at work on the days following.” Hine*

### Ongoing support and communication

Continuous support and communication with research nurses provided participants with the information and reassurance they needed to adhere to their treatment regimen, particularly when transitioning to SCIP. Wiremu and Mere appreciated the research nurse’s regular contact.

> *“[The research nurse] reminds me when it is coming up, I am pretty s** at remembering.” Wiremu*
>
> *“[The research nurse] reminds me and gives me choices for the next appointment.” Mere (mother)*

### Pain management

The final theme was pain management. This theme highlights the trade-off between experiencing occasional discomfort with SCIP versus the more frequent pain associated with IM injections.

### Differing levels of pain for participants

Pain levels varied between participants, with some participants finding SCIP more painful initially, while others reported less pain compared to the monthly IM injections.

> *“The one on my tummy hurts a lot less because whenever it does get sore, they just stick anaesthetic in it so it numbs everything out.” Teuila.*

Participants who experienced pain developed self-care methods to manage their/their child’s pain as they get more familiar with the procedure:

> *“Having a plan, Panadol and keeping him home for the day. He will rest for the whole day, and then he’s good to go the next day.” Kiri (mother)*
>
> “*For me I usually have a shower two hours after.” Wiremu*
>
> *“It hurts for a couple of days, but it’s way better than the injection trust me.” Teuila*

### Pain trade-off with fewer injections

While some participants experienced more pain with SCIP compared to IM injections, they found the trade-off worthwhile, as fewer injections and reduced interruptions to their daily lives made it a preferable choice. Mum’s Kiri and Mele know that it might be painful for their children, but it is worth it, knowing they only need SCIP every three months:

> *“So it’s a bit more painful after the injection, but they don’t have to worry about it every month. I’d rather they take the pain once every three months than deal with the monthly pain.” Kiri (mother)*
>
> *“The injections may hurt more with SCIP, but I’d rather go through a little more pain and not have to do it every month. It’s worth it for the freedom of fewer injections.” Mele*

Teuila, Hine and Ari reflect that they would rather put up with pain initially so they can forget about it for the next three months:

> *“It’s sore for a bit longer, but once it goes away, I know I have three months to not worry about it. I’d take that any day over the monthly one.” Hine*
>
> *“For first week the three-monthly one was worse, but I’d still prefer it over the monthly one. I just put up with it for a few days and it’s worth it.” Ari*

## Discussion

This study provides valuable insights into the lived experiences of Māori and Pacific Peoples with ARF who have been on SCIP for extended periods of time, along with the experiences of their whānau. Participants reported significant improvements in their quality of life with the transition from monthly IM injections to the less frequent SCIP treatments. These findings align with existing research highlighting that reduced frequency of injections can improve treatment adherence and reduce the physical and emotional burdens associated with treatment [13]. Participants and their whānau appreciated flexibility of the research nurses, which allowed for easier scheduling and smoother incorporation of treatment into their daily lives. This reflects the growing body of literature supporting the importance of whānau-centered models of care, which emphasise flexibility and responsiveness to Māori and Pacific contexts [14].

Participants’ experiences with SCIP also highlight the importance of strong patient – healthcare provider relationships, which is crucial for people who have disengaged due to past negative experiences within the healthcare system. The theme of Whakawhanaungatanga – building trust and open communication – was vital to empowering participants to transition from IM BPG to SCIP and ensure adherence to the new treatment regimen [15]. A positive relationship with research nurses was central to participants’ trust and confidence in their care. Aligning with studies that stress the role of continuous, supportive healthcare relationships in promoting better patient outcomes [16]. Furthermore, the reduction in emotional strain due to fewer injections and the ability to engage in normal life activities supports existing research that links treatment frequency with improved mental well-being [13].

In terms of pain management, participants reported varying levels of pain following the SCIP procedure, but all participants found the trade-off of less frequent injections worthwhile. Some noted initial pain and bruising following the injection, but the ability to manage these side effects with self-care strategies like rest and pain relief provided comfort. This reflects the broader literature on chronic treatment regimens, where patients often accept occasional discomfort in exchange for fewer treatments and less frequent medical visits [17].

The findings from this study are highly relevant for practice and policy, especially in developing culturally responsive healthcare models for Māori and Pacific communities. The success of SCIP in improving participants’ quality of life and treatment adherence supports its continued use in the Wellington region and suggests potential for expansion into other regions. The study also highlights the need for tailored interventions that respect cultural values, such as those embedded in Māori and Pacific cantered approaches to healthcare, ensuring that treatment plans are not only clinically effective but also culturally safe and responsive.

One of the key strengths of this study is that all participants had been on SCIP for at least four injections, with some having used SCIP for over two years. This provided rich, longitudinal insights into the sustained impact of SCIP on both health outcomes and quality of life. Moreover, the study included participants who had prior experience with IM injections, enabling direct comparisons between the two treatment regimens, further enriching the findings. However, some participants who had been recently diagnosed with ARF had been on SCIP for a longer period than IM BPG, which may have limited the ability to make a direct comparison. In addition, as the recruitment process relied on research nurses to recruit participants may have been more likely to be engaged with healthcare services. Finally, some participants were quieter with their whānau taking a more prominent role in the interview process, which may have affected the depth of their personal reflections.

While this study provides valuable insights into SCIP’s impact on patients’ lives, there is a need for future research to assess its broader implementation as a standard treatment across NZ. Research should focus on the feasibility of expanding SCIP into regions with alternative models of care, as well as areas with limited access to primary care and community support. Additionally, future studies could explore the barriers and facilitators to SCIP implementation in other countries to evaluate its global success.

## Conclusion

This study highlights the benefits of SCIP in reducing physical and emotional burdens, which in turn enhances the quality of life for Māori and Pacific participants with ARF. The findings emphasise the importance of culturally responsive, whānau-centred models of care. SCIP has the potential to improve adherence to secondary prophylaxis, with the potential to be offered as a standard treatment, improving health outcomes across NZ and internationally.

## Data Availability

Data sharing would need approval from indigenous governance groups. Data is verbatium interviews with participants.

## Acknowledgments

We thank the participants for their time and insights, we also thank the healthcare workers who have supported this study from Kenepuru, Hutt and Wellington Hospitals.

